# The acoustic quality and health in urban environments (SALVE) project: Study design, rationale and methodology

**DOI:** 10.1101/2021.02.23.21252275

**Authors:** Timo Haselhoff, Bryce Lawrence, Jonas Hornberg, Salman Ahmed, Robynne Sutcliffe, Dietwald Gruehn, Susanne Moebus

## Abstract

Sound pressure levels expressed in variations of decibel (dB) formulations are a common approach to describe the urban acoustic environment (AE). In recent years, different approaches gained traction to describe the urban AE, like the soundscape ecology approach, which focuses on the natural environment. To determine the feasibility of applying this approach to cities, a comprehensive dataset of high-quality sound recordings with high spatial and temporal resolution is essential.

The *acoustic quality and health in urban environments* (SALVE) project aims to establish a spatially and temporally high-resolution dataset of the urban AE using land use categories. Since 2019, we assess the AE at selected places in the densely populated city of Bochum, Germany. For a high temporal resolution, we used automatic devices at 52 locations that recorded every 26 minutes for three minutes. For a high spatial resolution, we used manual devices to perform a five-minute recording four times a year at 730 selected locations. Altogether, we ended up with 1,500,493 minutes of sound recordings.

Aim here is to outline our sampling design, methods used, and applied quality procedures in order to achieve a well-defined and high quality dataset presented for further scientific analysis. To the best of our knowledge, this represents one of the most extensive datasets currently available, which will provide a comprehensive database for future in-depth analyses of the associations between the urban AE, urban fabric and human health.

**Highlights:**

1. A conceptual sampling framework for measuring the urban acoustic environment is given
2. One of the most extensive datasets of the urban acoustic environment is introduced
3. Experiences and results of the field work of the SALVE-Project are presented

## 1 Introduction

The urban acoustic environment (AE) is mainly examined in terms of noise, operationalized as sound pressure level (SPL) and reported mostly as A-weighted equivalent continuous SPL (L_Aeq_), or averaged sound levels (L_day_, L_n_, L_dn_) [1]. Numerous studies including the comprehensive report of the European Environment Agency have shown the impact of noise on human health [2-11]. However, as the AE can be defined as “the sound from all sound sources modified by the environment” [12], it is likely that SPLs are not sufficient to capture the complexity of the whole AE. For example, Aletta et al. [13] published results of a systematic review assessing associations between positive health-related effects and perceptual soundscape constructs. Their findings suggest that there are more properties of the AE beyond noise that might have an impact on human health.

The soundscape approach allows the analysis of perception-based impacts of the urban AE on human health [14]. However, comprehensive studies demonstrating effects of urban soundscapes on health are still missing. A research field that examines sounds across a variety of different landscapes is the emerging science of soundscape ecology. Through a combination of expertise from spatial and acoustic ecology as well as bio- and psychoacoustics “coupled natural-human dynamics across different spatial and temporal scales” [15] can be described. Here, the use of multiple indices that take frequency and amplitude modulation into account is promoted to describe different properties of the AE [16].

So far, most studies analyzing the urban AE mainly focus on selected areas, like parks, green spaces, recreation areas or specific open spaces [17]. However, as the highly varying properties of the built environment are considered as one of the main influences on (perceived) sounds [14], further data is needed to describe the AE of all types of urban areas, including residential spaces, industrial zones or major and heavily trafficked roads. Previous studies that used spatial stratification, e.g. road type, grid method, land use and examined temporal factors, e.g. length of recording, number of samples, focused exclusively on noise indices [18-21]. Therefore, an essential requirement for the application of the soundscape ecology or perception-based approaches is the availability of comprehensive data in urban environments that considers the high temporal and spatial variability of the urban AE. The *acoustic quality and health in urban environments* (SALVE) study aims to establish a spatial and temporal high-resolution dataset of the urban AE using land use categories. The objective of this paper is to present the study design of SALVE including sampling design, methods used and quality assessment procedures as well as a description of the dataset generated.

## 2 Methods

### 2.1 SALVE study design

SALVE was launched at the end of 2018 as an interdisciplinary study, conducted by the Institute for Urban Public Health at the University Hospital Essen and the Department of Landscape Ecology and Landscape Planning at TU Dortmund University [22]. The main objective of SALVE is to describe the urban AE of the densely populated city of Bochum located in the polycentric metropolitan Ruhr Area, Germany. SALVE aims to establish a quality assured, spatially and temporally high-resolution dataset of the urban AE as a prerequisite to examine the association between the urban AE, the urban fabric and human health.

We pursued both a high spatial resolution to adequately capture the diversity of the built environment as well as a high temporal resolution to understand the daily, weekly, or seasonal variance of the AE [23]. Accordingly, we developed a sampling methodology to investigate the urban AE, considering characteristics of the built environment and available health data.

We applied a random sampling design that is used to apply statistical models to infer the target population. Main prerequisites for this are: (i) the selection probability must be known beforehand, (ii) and the probability of selection for each unit must be > 0 [24]. To satisfy these conditions, many surveys use sampling frames of the target population. As there is no frame of all possible built environments, we needed a way to approximate this. Additionally, due to limited resources, we needed a way to allocate our AE measuring devices efficiently. In the following, we describe the developed sampling framework, results, and limitations in detail.

### 2.2 Sampling Framework

We defined the target population as the AE of a subset of the urban environment in Bochum. As sounds are influenced by the built environment and this itself is extremely diverse, it must be approximated. We used these approximations as the sampling (or observation) unit. To depict the sound of the entire city, every unit should have a selection probability > 0 [24].

In SALVE, the urban environment was approximated by the land use (LU) defined by the “Regionalverband Ruhr” in 2015. The LU categorizes all land uses contiguously in the Ruhr Area into their respective designated residential, commercial, industrial, institutional or open space land uses [25]. As we were interested to analyze the association between the AE and health, we limited our target population to the LU categories that contained a residential component (‘Wohngebiet’). Also, the LU-Categories had to be included in a 600-meter buffer around residences of participants of the Heinz Nixdorf Recall (HNR) health study. The HNR is an ongoing population-based prospective cohort study with participants randomly selected from the lists of the *Office of Residence Registration*, including the city of Bochum [26]. For the city of Bochum, health data is available for 837 inhabitants. Additionally, we recorded the AE in the two most common forest types and designed public open spaces as a counterpoint. The three largest populations by category of urban green were chosen from the LU-Categories of Bochum. The resulting number of distinct areas for each LU-Category is given in table 1.

**Table 1:** Number of LU-Categories in Bochum and sampled LU-Areas for each procedure.

| LU-Category<sup>1</sup> | Bochum<sup>2</sup> | 600 m Buffer<sup>3</sup> | DAP<sup>4</sup> | AAP<sub>24</sub><sup>5</sup> | AAP<sub>4</sub><sup>6</sup> | BP<sup>7</sup> |
| --- | --- | --- | --- | --- | --- | --- |
| Residential areas, up to 3 floors | 4,245 | 4,155 | 94 | 3 | 3 | 6 |
| Residential areas, up to 5 floors | 2,824 | 2,807 | 93 | 3 | 3 | 6 |
| Residential areas, over 5 floors | 202 | 202 | 66 | 3 | 4 | 7 |
| Commercial mixed Use | 1,018 | 1,005 | 88 | 3 | 3 | 6 |
| Mixed industrial / residential areas | 1,273 | 1,169 | 90 | 3 | 4 | 7 |
| Public and institutional facilities | 239 | 234 | 69 | 3 | 4 | 7 |
| Agricultural land with homesteads | 131 | 107 | 56 | 3 | 4 | 7 |
| Designed neighborhood parks | 1,402 | 1,390 | 90 | 1 | 3 | 4 |
| Deciduous forest | 133 | 117 | 57 | 1 | 0 | 1 |
| Mixed forest | 36 | 35 | 27 | 1 | 0 | 1 |
| Total: | 11,503 | 11,221 | 730 | 24 | 28 | 52 |

<sup>1</sup>LU-Category of the “Regionalverband Ruhr”, own translation; <sup>2</sup>number of LU-Areas in Bochum; <sup>3</sup>number of LU-Areas in a 600 m Buffer around the HNR participants; <sup>4</sup>number of sampled LU-Areas from the respective LU-Category for the DAP; <sup>5</sup>number of sampled LU-Areas from the respective LU-Category for the AAP<sub>24</sub>; <sup>6</sup>number of sampled LU-Areas from the respective LU-Category for the AAP<sub>4</sub>; <sup>7</sup>number of sampled LU-Areas from the respective LU-Category for the BP.

The AE was operationalized as physical properties of the AE measured by microphones. We aimed to consider the composition of the whole frequency spectrum to be able to calculate acoustic indices, cross applied from the field of ecoacoustics, that represent characteristics other than noise [16, 27, 28]. Simultaneously, this approach allows the investigation of direct physical effects of urban sounds on humans, independent from their perception. Nevertheless, as human perception is an important process that modulates the effect of the AE, we also carried out binaural measurements according to DIN ISO 12913-1:2018-02 [12].

We applied three main sampling approaches: (i) a spatially high-resolution sampling approach, using portable sound level meters for manual measurements that follow a direct aural procedure (DAP), (ii) a temporally high-resolution sampling approach, using stationary recording devices that allow an automated aural procedure (AAP) and (iii) a binaural recording procedure (BP) according to the DIN ISO 12913-1:2018-02. The DAP approach allows for an economically reasonable number of devices to achieve a high spatial resolution. However, this approach requires rather high personnel and logistic expense and provides only a temporal snapshot of the AE. The AAP is widely used in ecoacoustics and allows for temporally dense recordings (i.e. high temporal resolution). However, this approach requires numerous devices, which are in jeopardy of being stolen or damaged. The BP allows for subsequent psychoacoustic studies related to our observation units to conform to DIN ISO 12913-2. Details of the respective recording methods are described in chapter 2.3.

With the target population (AE of Bochum), the observation unit (selected LU-Categories) and the measuring procedures defined (DAP, AAP & BP), the sampling methods can be developed as described in the following.

#### 2.2.1 Sampling Approach

We used a stratified sampling approach, as this has several advantages over a simple random sampling: (i) Estimations will be more exact (e.g. avoiding bias), (ii) sampling costs can be reduced and (iii) the strata themselves can be examined [24, 29]. In our case, the strata are based on the LU-Categories presuming that they differ in their AE because of the different properties of their urban and peri-urban environment. Accordingly, the primary sampling unit was the LU-Category, the secondary sampling unit the distinct LU-Area (i.e. the specific parcel in the city).

#### 2.2.2 DAP Sampling

The number of desired samples from each stratum was calculated using the formula:

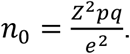

We corrected the results for finite populations, if necessary. Here, Z is defined by the desired α-level, *e* is the margin of error and *p* the proportion of the population with a given attribute. *q* equals *1-p* [30]. To use our resources efficiently, we defined an *α*-level of 5%, a margin of error of ± 10% and assumed maximum variability (*p* = 0.5) for the DAP.

In total, 730 LU-Areas were randomly sampled from their respective LU-Category. We assigned each area a random measurement point, using the ArcGIS add-in from NOAA [31]. Simultaneously, this tool allows the specification of the minimum distance between measurement points, which we set to 500 meters. Each recording point should be sampled four times (once every season). Thus it was estimated that 2920 DAP measurements had to be performed.

#### 2.2.3 AAP Sampling

Based on the limitation of 28 available automatic devices, we located three in each of the seven strata of interest plus one in each urban green category. The 24 devices are deployed for 365 days. We will refer to these devices in the following as AAP_24_. Additionally, we used four devices for explorative research interests. These devices are deployed for six weeks each and rotated to another location afterwards. These will be referred to as AAP_4_. This approach summarizes to 52 (24 + 28) sampled LU-Areas. The respective measurement points represent a convenient sample. The placement of the automatic devices was done in consultation with the city of Bochum. Using the tree cadaster of the city of Bochum [32], which lists trees on public and private ground, an available tree was selected within the LU-Area of all 52 sample locations.

#### 2.2.4 BP Sampling

The binaural recordings for the soundscape approach followed the AAP sampling and the recordings should be made once each season at the AAD_24_ and at each new sample location of the AAD_4_.

### 2.3 Measurement Procedures

#### 2.3.1 DAP Recordings

To organize the DAP sound recordings effectively, clustering of the 730 DAP sample points was performed in ArcGIS (10.5) with *Grouping Analysis* (*K_Nearest Neighbors*) to build 35 DAP point cluster. In order to optimize the travel times, we built a network dataset in ArcGIS from multilevel roadways then used the *New Route* function to created optimized routes for each cluster. This allowed the field staff to record five minutes at all points of one cluster in one day, assuming five minutes to prepare the device and an average walking speed of five km/h. As a result, 140 field days were necessary to measure all 730 sampled areas four times. The field days were distributed evenly over working days for 52 weeks. Days of the week on which measurements were to be taken were randomized to prevent selection bias. All measurements were scheduled between 9 a.m. and 5 p.m. As not all pre-defined sampling points were accessible (e.g. because of points “on” buildings, private places, construction work etc.), the field staff was advised to measure as near as possible to the pre-defined sampled points. If the new sampling point deviated by more than five meters from the pre-defined point and/or changed to an adjacent LU-Area, the new sampling point was documented in the field map and later updated in ArcGis. Thus, it was ensured that the follow-up measurements could be performed at the same place.

The DAP recording was done by mounting the microphone on a stand at around 1.65 m height, reflecting the approximated average height of a human ear [12]. The five-minute audio recording was carried out, using the NTi XL2 sound recording device with the M2230 omni-directional microphone [33]. When coupled with the MA 2230 microphone/preamp, the XL2 device was calibrated to meet the following IEC standards: IEC 61672:2013, IEC 61672:2003, IEC 61260:2014, IEC 61260:2003, IEC 60651, IEC 60804. The mono-recording was sampled at 48,000 Hz and with 24-bit depth. All files were stored in the WAVE format. Concurrently with each DAP measurement; weather-data was recorded in order to consider weather effects. Temperature, wind speed, humidity and atmospheric pressure were measured using the Skywatch Windoo 3 [34]. As we experienced that the Windoo device did not work all the time properly, the field staff was advised to use data from *weather*.*com* as a proxy and to document this in the field report. The fieldwork was paused or postponed in case of heavy rain to avoid biasing sounds and damage to the microphones.

#### 2.3.2 AAP Recordings

Upon each tree chosen from the tree cadaster, a Wildlife Acoustics SM4 Acoustic Recorder with a SMM-A2 microphone was mounted [35] at a height of approx. 1.65 m. The devices were configured to carry out a 3-minute measurement every 26 minutes, so we end up with a minimum of two recordings per hour of day. The mono-recordings were sampled at 44,100 Hz with a bit-depth of 16. The data was also stored as WAVE.

The AAP_24_ were set to record for 365 days straight and each of the AAP_4_ for six weeks, eight times the year. The devices needed battery and memory card replacements every six to eight weeks, resulting in eight maintenance days for AAP_24_ for one year. The same maintenance frequency applied to the AAP_4_. For all AAP, no weather data was recorded directly at the device.

#### 2.3.3 Binaural Recordings

Binaural recordings were planned once each season at the AAD_24_ and at each new sample location of the AAD_4_, resulting in an estimated number of 124 (24•4+28) recordings. For the binaural recordings, a 3Dio FS Pro II [36] with a ZOOM H4nPro stereo recording device [37] was used. The stereo-recordings had a length of five minutes, a sampling rate of 44,100 Hz and a bit depth of 24. Additionally, a 170 degree panorama picture was created for each recording location.

In total, 2 920 DAP-, 438 000 AAP_24_-, 58 800 AAP_4_- and 124 binaural-recordings were estimated by the developed sampling framework.

## 3 Results

DAP measurements were carried out from 18.03.2019 to 19.03.2020. AAP_24_ measurements since 06.05.2019 and AAP_4_ measurements since 01.08.2019. In total the recorded sound data of the SALVE project reaches 1,500,493 minutes, or 1042 days of continuous recording respectively. 1 % of them were recorded using the DAP, 83 % using the AAP_24_, 16% using the AAP_4_ and 0,04 % using the binaural procedure (table 2).

**Table 2:**
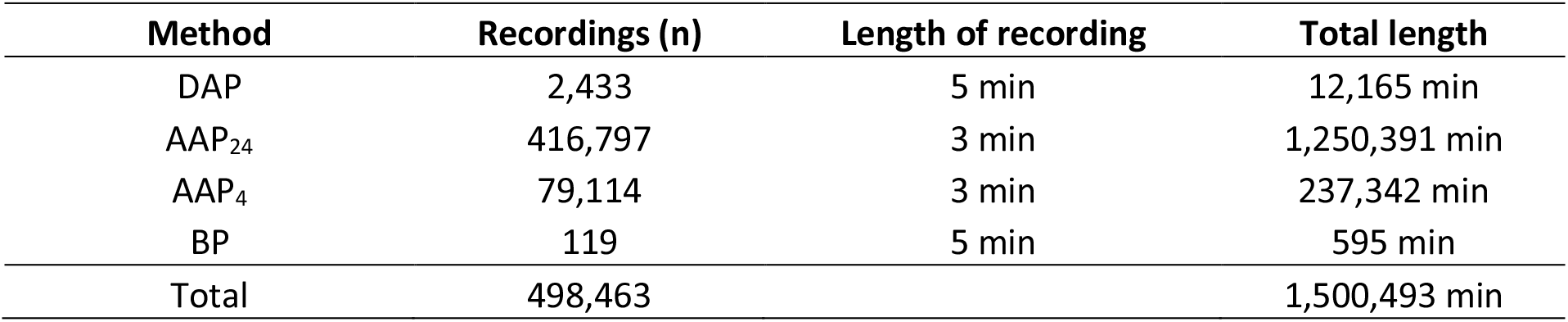
Quantity of recordings by procedure In total the dataset has a size of 7.44 Terabyte (DAP = 100 GB; AAP24 = 6.16 TB, AAP4 = 1.17 TB, BP = 5.4 GB)

The final dataset contains fewer measurements than estimated above due to common field measurement disruptions, such as inclement weather conditions, technical equipment failures, and personnel error. Problems of the DAP were mainly caused by an initially wrong factory calibration of one microphone and cable shorts. As the field collection started in early spring, most of the disruptions occurred in this time (Appendix A.1). Initially not considered LU-Categories have been included due to deviations from the pre-defined measurement points (mean = 17.2 m). Reasons were mainly that predefined locations were placed on private property, commercial or industrial locations. Further reasons for changing or excluding pre-definded measurement points included pedestrians talking directly into the microphone, measurements outside the desired timespan or individual mistakes in locating the device. Regarding the weather data, all Windoo devices had several malfunctions, resulting in missing full weather data for 14 % (n = 347) of the recordings. By means of various organizational and logistical modifications the DAP measurements are not evenly distributed over the working days (Appendix A.2). Most measurements were deployed in the given time-span from 9 a.m. to 5 p.m. with only 72 recordings outside the planned interval. The frequencies of the recordings by daytimes are depicted in Appendix A.3. The distribution with peaks at 11 and 12 a.m. and lowest number at 9 a.m. and 4 p.m. are justified by the given routes per cluster.

The AAP_24_ devices were deployed on 06.05.2019. One device was unknowingly damaged shortly after deployment, which we only noticed well into the measurement period so that a replacement was no longer reasonable. Hence, only 23 devices covering the same recording period are available for data analysis. Other reasons for recording dropouts have occurred for the following reasons: one device could not be put into operation for six weeks, two devices ran into battery problems and one device was destroyed with an axe and replaced after a two month gap. For an overview of all recording periods, see Appendix B.3. The AAP_4_ devices were first deployed on 01.08.2019 changing locations six times until 06.07.2020. One issue with the locks we used to mount the devices resulted in leaving the devices in the same place for an additional six weeks. Thus, the overall number of measurement exceeds the estimated one with 79,114 files recorded. Given the limitation of using trees as AAP device mounting location we had mount AAP devices a small distance away from the target LU-Area in some cases. The target and the actual measured LU-Category for all AAPs can be found in Appendix B.

The BP was mostly carried out as planned, however, we measured twice in the spring for the AAD_24_ and thus skipped the summer recordings. Therefore, we ended up with the estimated number of recordings, but with different periods measured. The recordings at the AAD_4_ went mostly as planned, but five recordings were missed out due to reasons describe above (Appendix B.2).

The sound measurements were mostly received positively by the citizens. One reason might have been, that information flyers were placed in every mailbox surrounding the devices location and a sticker for further information was put on each device. Nevertheless, two AAP devices had to be moved by about 300 meters due to resident complaint. Also, the police was called by suspicious residents three times during DAP field days. However, the team member informed the police and every time the police officers saw no issues with the project, so that the measurements could be continued.

**Figure 1:**
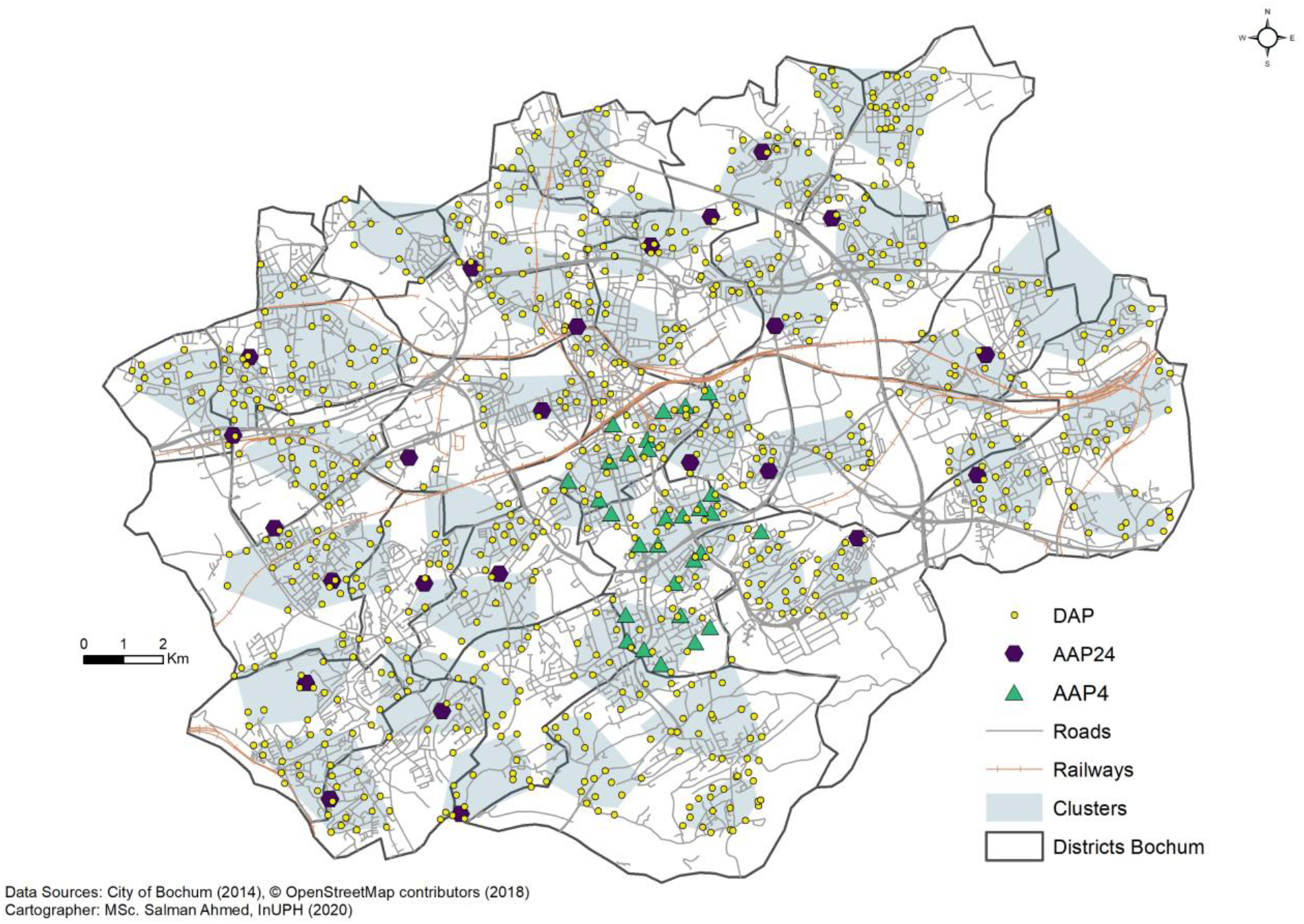
Measured points by recording procedure in Bochum, Germany.

## 3.1 Quality assurance

To ensure the quality of the measurements, we implemented an accompanying quality assurance. Prior to the fieldwork, main study elements, like implementation, application, and evaluation of the study were documented in a detailed and binding study protocol. Rules for and comments on the conduct of the data collection were fixed in a written form and made available to the field team. Changes during the field phase were documented as amendments. The study personnel involved in the field study were skilled and trained prior to the beginning of the fieldwork. The training comprised a theoretical introduction to the study objective, training the use of devices and field procedures, followed by one field day with an experienced guide. Monthly team meetings ensured a regular exchange of experiences and an adjustment of methods, if necessary. Calibrations for all devices were checked beforehand and their settings were streamlined. The collected data was comprehensively checked for plausibility, using descriptive statistics on all measured variables. Large deviations or potentially implausible data were discussed between all team members and results were documented accordingly. Necessary changes were saved in a new dataset.

## 4 Discussion

In SALVE, a methodology was developed and applied to assess the urban AE in the City of Bochum, taking into account the built environment. The resultant dataset is proof of concept of the sampling and data collection procedure and created a dataset to further understand the urban AE. The measurement techniques for each procedure were outlined as well as their applications in the field. Many of these were similar to the applications in ecoacoustics, but additional complications, mostly regarding fieldwork in the urban environment, had to be solved. The suggested methods can be easily adapted in surveys of the AE in other cities. Nevertheless, future analysis of the recorded sound data of SALVE must prove if the framework is appropriate for representing the AE in dependence to the built environment.

Altogether, the data of the SALVE project represents a disproportionally stratified random sample of LU-Areas in a 600-meter buffer around participants of the HNR for the DAP and a convenient sample with a temporal resolution of approx. two three minute recordings per hour for each device over one year for the AAP. In addition, the BP generated 119 binaural recordings over one year.

Although the DAP provided a high spatial resolution, there were difficulties in the field, resulting in a possibly biased dataset, if sounds differ systematically by working day and/or daytime. Still the DAP data provides the possibility to investigate the AE in a high spatial resolution. The AAP on the other hand had fewer issues: the resulting recordings allow for detailed analysis with a high temporal but low spatial resolution. The binaural recordings and their respective panorama pictures allow for analysis using the perceptual soundscape approach. All data will be made available for scientific research.

### 4.1 Strengths and limitations

One of the strengths of SALVE is the systematic approach to capture the AE of a wide range of types of urban areas taking into account possible temporal variances. A further strength is the implementation of a rigorous quality management procedure, including the application of a study protocol, qualification and training of the field staff, plausibility checks of assessed sound recordings and all measured variables. Furthermore, linking the sound data to results of a population-based health study will likely bring new insights of effects of the urban AE on humans. For the future, the recorded sound data will allow a great variety of analysis. Due to the length, the sampling frequency, and the uncompromised file-format of the sound data, a wide range of sound information is available. Especially the high spatial and temporal resolution enables detailed investigations of the properties of the AE that opens up the possibilities e.g. of using machine learning approaches. As neither the DAP nor the AAP focusses solely on one dimension, inter and intra comparisons of the temporal and spatial structure of urban sounds are possible.

Some limitations of the study deserve special mention. The sampling design aims at recording the AE of Bochum as efficiently as possible. However, LU-Categories were restricted to the residentially related land uses within a 600-meter buffer of the participants of the HNR study. Therefore, not all LU-Categories were taken into account. Although the HNR is a simple random sample of the population between 45 and 75 years [26] and our DAP sample therefore still is a random sample of a random sample (ignoring panel-effects), it is not a random sample of the urban environment of Bochum anymore. Secondly, regarding the DAP not all working days were sampled equally which leads to an oversampling on Wednesdays and Thursdays and undersampling of Mondays and Fridays. However, since we also have AAD recordings available with records during the whole week, the magnitude of this oversampling can likely be determined. Next steps are to check if the LU is an appropriate approximation for the built environment to describe the pattern of the urban AE. For instance, if the AE inside the same LU-Category is not more similar than the ones between the different LU-Categories, a stratification by LU can be hardly justified for future studies. The same is true for temporal differences.

## 5 Conclusion

With SALVE we achieved a longitudinal set of recordings systematically gathered from the urban environment with both automated and manual procedures, which is so far one of the largest AE datasets ever recorded. This dataset can be used for a wide array of upcoming studies. The innovative element of this study is that it couples the field of epidemiology with the field of ecoacoustics and urban planning, thereby treating urban land use as so called “human habitat patches” and seeks to use sound as an indicator for habitat quality – similar to the use of sound in ecoacoustics as measures of non-human animal habitat quality. Our aim is to characterize the urban environment taking into account the extent of differences and similarities between recordings of the AE within different urban land use types. This approach will lead to an understanding of the nature and complexity of sounds in the human dominated urban fabric.

Overall, we aim to address questions especially regarding associations between the built environment and the AE, associations between human health and the AE, connection of physical sound measurements and human perception and a methodology for appropriate sound sampling to represent the sounds of the urban environment.

## Supporting information

Appendix A: DAP Data

Appendix B: AAP Data

## Data Availability

All data from the SALVE Project will be made available for scientific research.

## Abbreviations

AAP: Automated aural procedure
AAP_24_: 24 stationary devices of the automated aural procedure
AAP_4_: Four rotation devices of the automated aural procedure
AE: Acoustic environment
BP: Binaural procedure
DAP: Direct aural procedure
LU: Land use defined by the Regionalverband Ruhr in 2015

