## Appendix A: DAP Data for "The acoustic quality and health in urban environments (SALVE) project: Study design, rationale and methodology"

### A Appendix: Additional information on DAP recordings

A.1: Summary of DAP-Recordings by LU-Category (n)

| LU (Code) | Measured LU-Category | Spring | Summer | Fall | Winter | Total |
|---|---|---|---|---|---|---|
| 10 | Residential areas, up to 3 floors | 57 | 80 | 91 | 89 | 317 |
| 20 | Residential areas, up to 5 floors | 59 | 69 | 81 | 82 | 291 |
| 30 | Residential areas, over 5 floors | 31 | 53 | 54 | 55 | 193 |
| 40 | Commercial mixed use | 49 | 63 | 68 | 74 | 254 |
| 51 | Commercial area (facilities / residential areas) | 31 | 51 | 55 | 59 | 196 |
| 52 | Commercial area (storage spaces) | 7 | 9 | 12 | 11 | 39 |
| 53 | Commercial area (open / reserve areas) | 1 | 1 | 1 | 1 | 4 |
| 54 | Commercial area (parking space) | 2 | 5 | 6 | 6 | 19 |
| 84 | Public and institutional facilities (kindergartens, after-school care, youth and elderly homes / housing estates) | 29 | 44 | 54 | 50 | 177 |
| 85 | Public and institutional facilities (churches and parish halls, monasteries) | 0 | 2 | 0 | 2 | 4 |
| 86 | Public and institutional facilities (police, fire department, rescue stations, bunkers) | 0 | 1 | 1 | 1 | 3 |
| 91 | Agricultural land with homesteads (buildings and facilities) | 23 | 29 | 32 | 33 | 117 |
| 93 | Agricultural land with homesteads (other areas) | 0 | 1 | 1 | 1 | 3 |
| 140 | Major roads and main roads | 8 | 3 | 8 | 7 | 26 |
| 151 | Residential and access roads | 22 | 29 | 34 | 34 | 119 |
| 152 | Other paths / roads | 1 | 2 | 2 | 2 | 7 |
| 160 | Pedestrian zones | 1 | 1 | 1 | 1 | 4 |
| 171 | Parking spaces | 2 | 4 | 4 | 4 | 14 |
| 172 | Parking garages | 1 | 1 | 1 | 1 | 4 |
| 174 | Garage yards in residential and mixed use areas | 2 | 2 | 4 | 4 | 12 |
| 271 | Designed neighbourhood parks in residential and mixed use areas | 46 | 59 | 73 | 74 | 252 |
| 272 | Parks and gardens (municipal parks, botanical gardens, zoo) | 1 | 1 | 1 | 1 | 4 |
| 273 | Informal neighbourhood green spaces | 4 | 3 | 5 | 5 | 17 |
| 291 | Small gardens / open spaces in residential and mixed use areas | 1 | 2 | 2 | 2 | 7 |
| 321 | Roadside vegetation and tree alley | 2 | 3 | 3 | 3 | 11 |
| 361 | Meadows and pastures | 6 | 5 | 6 | 6 | 23 |
| 370 | Farmland | 1 | 3 | 3 | 3 | 10 |
| 400 | Deciduous forest | 39 | 45 | 54 | 52 | 190 |
| 410 | Coniferous forest | 1 | 1 | 1 | 1 | 4 |
| 420 | Mixed forest | 23 | 26 | 24 | 26 | 99 |
| 431 | Woods | 3 | 3 | 3 | 4 | 13 |
| | **Total** | **453** | **601** | **685** | **694** | **2,433** |

#### A.2: Summary of DAP-Recordings by working day

| Working day | n |
|---|---|
| Monday | 322 |
| Tuesday | 337 |
| Wednesday | 834 |
| Thursday | 636 |
| Friday | 304 |
| **Total** | **2,433** |

#### A.3: Summary of DAP-Recordings by hour of day

| Time period | n |
|---|---|
| 09:00 - 10:00 | 137 |
| 10:00 - 11:00 | 377 |
| 11:00 - 12:00 | 480 |
| 12:00 - 13:00 | 475 |
| 13:00 - 14:00 | 390 |
| 14:00 - 15:00 | 291 |
| 15:00 - 16:00 | 192 |
| 16:00 - 17:00 | 91 |
| **Total** | **2,429** |
