## Appendix B: AAP Data for "The acoustic quality and health in urban environments (SALVE) project: Study design, rationale and methodology"

### B Appendix: Additional information on AAP recordings

#### B.1: Target LU and measured LU of the $AAP_{24}$

| Device ID | Target LU | Target LU (Code) | Measured LU | Measured LU (Code) | Distance to target LU in meter |
|---|---|---|---|---|---|
| 1 | Residential areas, up to 3 floors | 10 | Small gardens / open spaces in residential and mixed use areas | 291 | < 5 |
| 2 | Residential areas, up to 3 floors | 10 | Small gardens / open spaces in residential and mixed use areas | 291 | < 5 |
| 3 | Residential areas, up to 3 floors | 10 | Residential areas, up to 3 floors | 10 | 0 |
| 4 | Residential areas, up to 5 floors | 20 | Informal neighbourhood green spaces | 273 | 5 |
| 5 | Residential areas, up to 5 floors | 20 | Tree groups and tree rows | 432 | 17 |
| 6 | Residential areas, up to 5 floors | 20 | Roadside vegetation and tree alley | 321 | 15 |
| 7 | Residential areas, over 5 floors | 30 | Garage yards in residential and mixed use areas | 174 | 10 |
| 8 | Residential areas, over 5 floors | 30 | Informal neighbourhood green spaces | 273 | 10 |
| 9 | Residential areas, over 5 floors | 30 | Designed neighbourhood parks in residential and mixed use areas | 271 | 28 |
| 10 | Public and institutional facilities (kindergartens, after-school care, youth and elderly homes / housing estates) | 84 | Residential and access roads | 151 | < 5 |
| 11 | Public and institutional facilities (kindergartens, after-school care, youth and elderly homes / housing estates) | 84 | Residential and access roads | 151 | 30 |
| 12 | Public and institutional facilities (kindergartens, after-school care, youth and elderly homes / housing estates) | 84 | Informal neighbourhood green spaces | 273 | 8 |
| 13 | Commercial area (facilities / residential areas) | 51 | Informal neighbourhood green spaces | 273 | 20 |
| 14 | Commercial area (facilities / residential areas) | 51 | Commercial area (facilities / residential areas) | 51 | 0 |
| 15 | Commercial area (facilities / residential areas) | 51 | Residential and access roads | 151 | 6 |
| 16 | Designed neighborhood parks in residential and mixed use areas | 271 | Sports and play facilities | 308 | < 5 |
| 17 | Agricultural land with homesteads (buildings and facilities) | 91 | Mixed forest | 420 | 108 |
| 18 | Agricultural land with homesteads (buildings and facilities) | 91 | Small gardens / open spaces in residential and mixed use areas | 291 | < 5 |
| 19 | Agricultural land with homesteads (buildings and facilities) | 91 | Agricultural land with homesteads (buildings and facilities) | 91 | 0 |
| 20 | Deciduous forest | 400 | Deciduous forest | 400 | 0 |
| 21 | Commercial mixed use | 40 | Small gardens / open spaces in residential and mixed use areas | 291 | < 5 |
| 22 | Commercial mixed use | 40 | Residential and access roads | 151 | 40 |
| 23 | Commercial mixed use | 40 | Residential and access roads | 151 | < 5 |
| 24 | Mixed forest | 420 | Mixed forest | 420 | 0 |

*Distance < 5 is used, when the device was placed upon trees on the edge of the target LU-Area.*

#### B.2: Target LU and measured LU of the AAP$_4$

| Device ID | Target LU | Target LU (Code) | Measured LU | Measured LU (Code) | BP |
|---|---|---|---|---|---|
| 25 | Commercial mixed use | 40 | Informal neighbourhood green spaces | 273 | Yes |
| 26 | Residential areas, up to 5 floors | 20 | Parking spaces | 171 | Yes |
| 27 | Designed neighborhood parks in residential and mixed use areas | 271 | Major roads and main roads | 140 | Yes |
| 28 | Residential areas, up to 3 floors | 10 | Woods | 451 | Yes |
| 25 | Public and institutional facilities (kindergartens, after-school care, youth and elderly homes / housing estates) | 84 | Other paths / roads | 152 | No |
| 26 | Agricultural land with homesteads (buildings and facilities) | 91 | Woods | 431 | Yes |
| 27 | Commercial area (facilities / residential areas) | 51 | Commercial area (open / reserve areas) | 53 | Yes |
| 28 | Residential areas, over 5 floors | 30 | Public and institutional facilities (churches and parish halls, monasteries) | 85 | Yes |
| 25 | Designed neighborhood parks in residential and mixed use areas | 271 | Residential areas, up to 3 floors | 10 | Yes |
| 26 | Residential areas, up to 3 floors | 10 | Residential and access roads | 151 | Yes |
| 27 | Commercial mixed use | 40 | Residential and access roads | 151 | Yes |
| 28 | Residential areas, up to 5 floors | 20 | Public and institutional facilities (public and private educational institutions, libraries) | 83 | Yes |
| 25 | Public and institutional facilities (kindergartens, after-school care, youth and elderly homes / housing estates) | 84 | Roadside vegetation and tree alley | 321 | Yes |
| 26 | Agricultural land with homesteads (buildings and facilities) | 91 | Roadside vegetation and tree alley | 321 | Yes |
| 27 | Residential areas, over 5 floors | 30 | Cemeteries | 282 | Yes |
| 28 | Commercial area (facilities / residential areas) | 51 | Parking spaces | 171 | Yes |
| 25 | Residential areas, up to 3 floors | 10 | Residential areas, up to 3 floors | 10 | No |
| 26 | Designed neighborhood parks in residential and mixed use areas | 271 | Residential areas, up to 5 floors | 20 | No |
| 27 | Commercial mixed use | 40 | Parking spaces | 171 | No |
| 28 | Residential areas, up to 5 floors | 20 | Commercial mixed use | 40 | No |
| 25 | Public and institutional facilities (kindergartens, after-school care, youth and elderly homes / housing estates) | 84 | Residential and access roads | 151 | Yes |
| 26 | Agricultural land with homesteads (buildings and facilities) | 91 | Woods | 431 | Yes |
| 27 | Residential areas, over 5 floors | 30 | Parks and gardens (municipal parks, botanical gardens, zoo) | 272 | Yes |
| 28 | Commercial area (facilities / residential areas) | 51 | Roadside vegetation and tree alley | 321 | Yes |
| 25 | Agricultural land with homesteads (buildings and facilities) | 91 | Informal neighbourhood green spaces | 273 | Yes |
| 26 | Residential areas, over 5 floors | 30 | Major roads and main roads | 140 | Yes |
| 27 | Commercial area (facilities / residential areas) | 51 | Informal neighbourhood green spaces | 273 | Yes |
| 28 | Public and institutional facilities (kindergartens, after-school care, youth and elderly homes / housing estates) | 84 | Major roads and main roads | 140 | Yes |

B.3: Recording period for all $AAP_{24}$

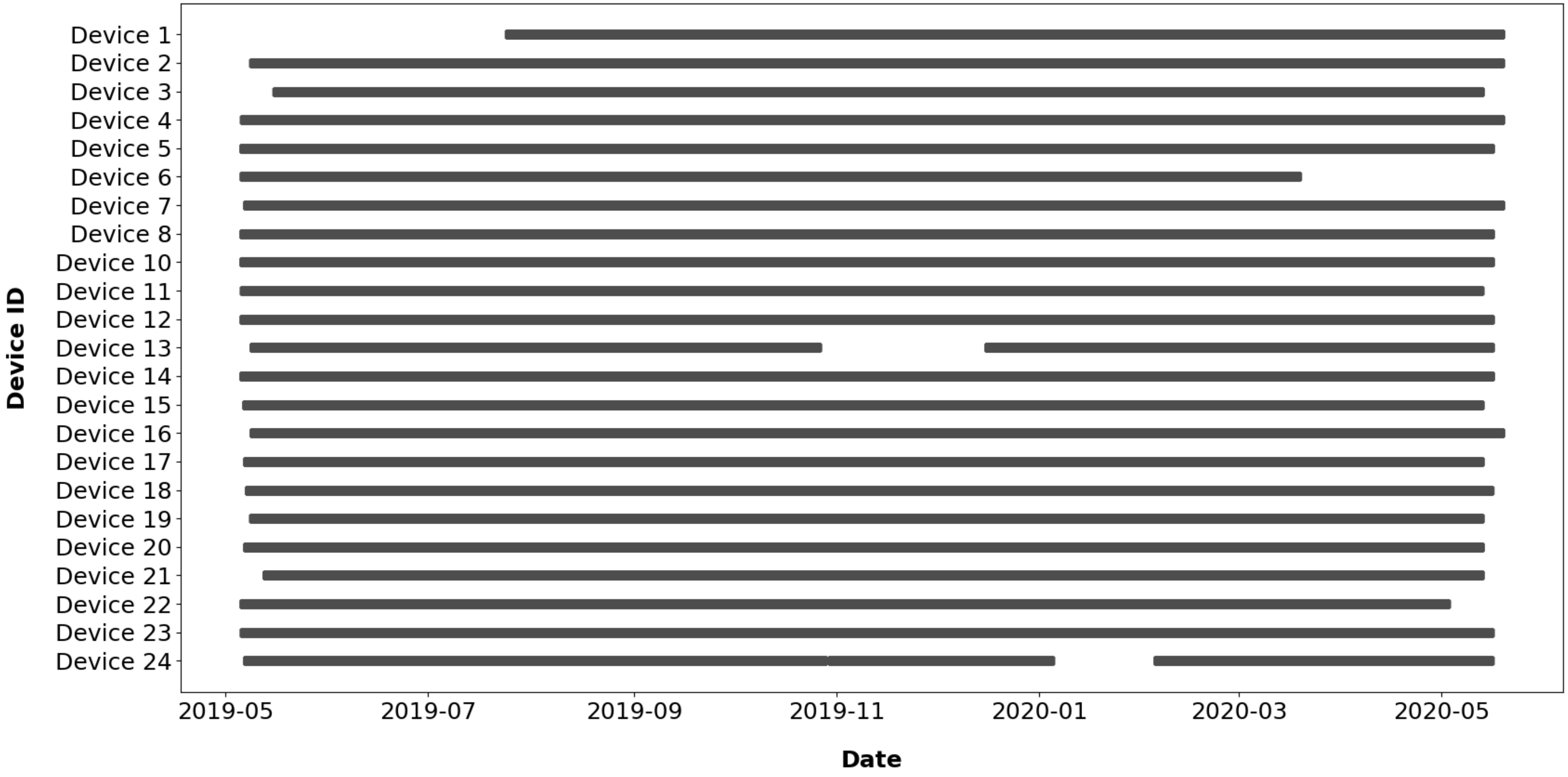
